# Predictors of mortality among post-COVID-19 discharged patients in Northern India

**DOI:** 10.1101/2023.03.06.23286877

**Authors:** Arjun, Basavaraj Jatteppanavar, Prasan Kumar Panda, Pathik Dhanger

## Abstract

**Background:** The one-year post-discharge all-cause mortality rate of COVID-19 disease is 7.87 % with the majority of patients’ readmission and mortality occurring within the first 30 days post-discharge.

**Objective:** Understanding predictors of mortality will help in prioritising patient care and preventive approaches.

**Methods:** Ours a single-centre unmatched case control study at a tertiary care centre in northern India, conducted from April 2020 to September 2022. The data was extracted retrospectively from the electronic hospital medical records of patients and by trained physicians using standardised data extraction sheet.

**Results:** A total of 184 patients were enrolled with 92 cases and 92 controls. The mean age of patients was 49.3 ± 17.53 years. The mortality group had a higher mean age (53.24 ± 18.53 yrs) as compared to the control group (45.37 ± 15.58 yrs) [p – 0.002]. Bivariate analysis revealed a significant difference in the two groups with respect to O2 saturation at admission [Case - 91.12 ± 12.49 %, control - 95.46 ± 5.01 %, p – 0.003); Maximum O2 flow rate [L/min] (Case – 11.01 ± 22.2, Control – 6.41 ± 13.31, P – 0.04); ICU need (p – 0.005), Cancer (p – 0.001), O2 need at discharge (p – 0.001) and AKI (p – 0.007). On multiple regression analysis, Cancer (aOR-2.469; 95% CI-1.183-5.150, p-0.016), ICU admission (aOR- 2.446; 95% CI-1.212-4.938, p- 0.013), Oxygen at discharge (aOR- 2.340; 95% CI-0.971-5.640, p-0.0586) and Acute kidney injury (aOR- 5.6; 95% CI-2.351-13.370, p-0.00) only found to be significant.

**Conclusion:** Oxygen requirement at discharge (2.3 times), Malignancy (2.4 times), ICU admission (2.4 times), and Acute Kidney Injury (5.6 times) were risks of death among COVID-19-recovered patients, post discharge. The presence of these variables would warrant a close follow up for these patients in order to decrease post COVID mortality.

## Introduction

The global pandemic of severe acute respiratory syndrome coronavirus Cov-2 (COVID-19) has caused more than 6 million deaths (1) and India has been one of the most severely affected countries with over 458,000 deaths reported by November 1, 2021 (2). The increasing rates of vaccination across the globe has contributing to declining disease incidence, however the substantial mortality still exists (3). The emergence of new strains of the virus increases the likelihood of repeat surge of cases and deaths due to the disease. The predictors of morbidity and mortality have been well studied for Acute COVID-19 in hospitalized patients, but similar data regarding the post COVID-19 illness is still accumulating.

Outpatient post COVID-19 data suggests persistence of multiple symptoms in this vulnerable population and development of new cardiovascular and pulmonary complications (4). All cause 30 day mortality and 30-day readmission rate of Post COVID-19 patients discharged on home based oxygen therapy was reported as 1.3% and 7.5% respectively in a retrospective cohort study (5). These figures may differ significantly in an Indian setting as has been described in the literature for acute COVID-19 illness (6). Ethnicity, gender, comorbidities, severity of symptoms, amongst others, determined the mortality and morbidity of an acute COVID-19 illness (7).

Such understanding of the predictors helps in prioritizing patient care and preventive approaches. However, similar data regarding the post COVID-19 mortality has been scarce. There is limited data from India regarding the prevalence and predictors of post COVID-19 mortality. This single centre hospital-based study was done with an objective of identifying the predictors of mortality among symptomatic COVID-19 patients discharged from the hospital setting.

### Methodology

This was a single centre hospital-based unmatched case control study conducted at a tertiary care centre in Northern India. COVID-19 Real Time Reverse Transcriptase Polymerase Chain Reaction (RTPCR) positive patients who were discharged from the hospital during the study duration and died within a 3-month period after discharge, were considered as cases. COVID-19

RTPCR positive patients discharged from the hospital during the same time, and alive for the 3-month post discharge period were considered as controls. The study was conducted over a period of 16 months from April 2021 to September 2022. All the patients of COVID-19 illness over the age of 18 years, who were admitted in the hospital and died during follow up were recruited in the study. The controls were unmatched and were selected randomly from the hospital records randomly from the hospital records in the ratio of 1:1. All the information related to the controls and cases were extracted retrospectively from the medical records of patients.

Patient related information was extracted by trained physicians using standardized data extraction sheet. The data extracted included demographic variables like Age, Sex, Smoking, Alcohol consumption, comorbidities like hypertension, diabetes, heart disease, Mucor-mycosis, cancer, chronic kidney disease and heart disease. Vitals at the time of presentation-Heart rate, respiratory rate, blood pressure were extracted. Data regarding symptom duration before admission, duration of hospital stay, saturation at admission, maximum O2 requirement at admission, type of respiratory support (nasal prongs, face mask, non-rebreathing mask, high flow nasal cannula, non-invasive ventilation, invasive ventilation), duration of oxygen requirement in hospital and need for oxygen at discharge was collected. Imputation was not done for missing data. All the study variables were represented either as means or proportions and compared between cases and controls using chi square test or unpaired t-test wherever appropriate. Univariable and multivariable logistic regression was done to assess association between various predictor variables and deaths due to COVID-19. A p<0.05 was considered as significant. Ethical approval for the analysis was obtained from the Ethics Committee of the All-India Institute of Medical Sciences, Rishikesh and all participating institutes (CTRI/2020/08/027169).

## Results

A total of 184 patients were included in the study; 92 were cases and 92 were controls. The mean age of patients was 49.3 ± 17.53 years. 65.46% were males and 34.54% were females. Cough was the most common symptom at the time of admission (53.1%). Followed by dyspnea present in 31.44% of cases. The mean duration of symptom onset before presentation to the hospital was 4.41 ± 3.85 days and the mean hospital stay was 12.12 ± 10.61 days. 34.53 % patients required oxygen need at admission while 40.20 % required oxygen anytime during the hospital stay.

24.74 % of all patients required ICU care with 4.63% of all requiring vasopressor support. 4.63% received invasive ventilation while 8.24% received non-invasive ventilation. 10.81% of all patients required Oxygen at discharge. (Table. 1)

**TABLE 1.** Demographic and clinical characteristics of patients and controls

|  |  |  |  |  |
| --- | --- | --- | --- | --- |
| **Characteristics.** | **Total**<br>**(n = 194)** | **Cases**<br>**(n = 92)** | **Controls**<br>**(n = 92)** | **P-**<br>**value** |
| **Age (years)** | 49.3 ± 17.53 | 53.24 ± 18.53 | 45.37 ± 15.58 | **0.002** |
| **Sex** |  |  |  | 0.5 |
| **M** | 127 | 64 | 63 |  |
| **F** | 57 | 28 | 29 |  |
| **Symptom duration before admission (days)** | 4.41 ± 3.85 | 4.46 ± 4.43 | 4.35 ± 3.25 | 0.848 |
| **Duration of Hospital Stay (days)** | 12.12 ± 10.61 | 11.59 ± 11.10 | 12.65 ± 10.12 | 0.498 |
| **Saturation at admission (%)** | 93.27 ± 9.76 | 91.12 ± 12.49 | 95.46 ± 5.01 | **0.003** |
| **Max Flow (L/min)** | 8.55 ± 18.15 | 11.01 ± 22.2 | 6.41 ± 13.41 | **0.04** |
| **Duration of O2 in hospital (days)** | 4.88 ± 6.93 | 4.65 ± 6.84 | 5.08 ± 7.03 | 0.715 |
| **Duration of ICU stay (days)** | 4.04 ± 8.13 | 4.92 ± 9.16 | 3.31 ± 7.14 | 0.249 |
| **O2 Flow at discharge (L/min)** |  | 1.50 ± 4.12 | 0.03 ± 0.183 | **0.007** |
| **O2 need at admission** | 67 | 33 | 34 | 0.5 |
| **O2 need during admission** | 78 | 38 | 40 | 0.466 |
| **Vasopressor Need** | 9 | 7 | 2 | **0.077** |
| **ICU Need** | 48 | 31 | 6 | **0.005** |
| **Dyspnoea** | 61 | 44 | 17 | **0.001** |
| **Hypertension** | 49 | 24 | 25 | 0.536 |
| **Diabetes** | 43 | 22 | 21 | 0.45 |
| **Mucor** | 8 | 5 | 3 | 0.33 |
| **Cancer** | 26 | 20 | 6 | **0.001** |
| **Heart Disease** | 24 | 12 | 12 | 0.528 |
| **Chronic Kidney Disease** | 21 | 14 | 7 | **0.059** |
| **O2 need at discharge** | 21 | 19 | 2 | **0.001** |
| **AKI** | 13 | 11 | 2 | **0.007** |

On bivariate analysis, as compared to controls, the cases were more likely to be older (53.24 years vs 45.37 years), with co-morbidities (cancer – 21.73% vs 6.5%, chronic kidney disease – 15.21% vs 7.60%). Cases were more likely to have dyspnea (47.82% vs 18.47%), lower saturation (91.12% ± 12.49 vs 95.46 ± 5.01%) at the time of admission as compared to controls. During the duration of hospital stay, the cases were more likely to have higher oxygen flow rates (11.01 ± 22.2 L/min vs 6.41 ± 13.41 L/min), more need for ICU admission (33.69% vs 6.50%), development of AKI (11.95% vs 2.17%) and more incidence of vasopressor need (7.60% vs 2.17%) as compared to controls. Cases were more likely to have invasive ventilation (7.60% vs 2.17%) and non-invasive ventilation (10.86% vs 6.52%). At the time of discharge, the cases were more likely to require oxygen at discharge (20.60% vs 2.17%) as compared to controls. (Table 1)

On multivariable analysis, ICU need (aOR 2.44, 95% CI 1.212 – 4.938), O2 requirement at discharge (aOR 2.34, 95% CI 0.971 – 5.640), Cancer (aOR 2.46, 95% CI 1.18 – 5.15) and AKI during hospital stay (aOR 5.60m 95% CI 2.35 – 13.37) were associated with post COVID mortality. After adjusting for all variables, sex, comorbidities like hypertension and diabetes were not associated with increased post COVID mortality (Table 2)

**TABLE 2:** Association between demographic, clinical characteristics and mortality among COVID-19 patients discharged from the hospital

| **Variable** | **aOR** | **95% CI** | **p-value** |
| --- | --- | --- | --- |
| **Age** | 1.03 | 1.00 – 1.06 | **0.01** |
| **Sex** | 1.29 | 0.72 – 2.33 | 0.38 |
| **ICU need** | 2.44 | 1.21 – 4.93 | **0.01** |
| **Vasopressor need** | 0.94 | 0.25 – 3.41 | 0.92 |
| **Need for O2 at discharge** | 2.34 | 0.97 – 5.64 | **0.05** |
| **Acute Kidney Injury** | 5.60 | 2.35 – 13.3 | **0.01** |
| **Malignancy** | 2.46 | 1.18 – 5.15 | **0.01** |
| **Diabetes** | 1.17 | 0.56 – 2.41 | 0.66 |
| **Mucor** | 1.26 | 0.27 – 5.95 | 0.76 |
| **Hypertension** | 0.79 | 0.40 – 1.57 | 0.51 |

Using multiple regression analysis to estimate adjusted Odds Ratio (aOR) and 95% confidence interval (CI).

## Discussion

In this hospital-based control study of predictors of mortality in patients discharged from a hospital setting after COVID-19 illness, we found that older age, oxygen saturation at admission, maximum oxygen requirement during hospital stay, flow rate at discharge, ICU admission need during hospital stay, cancer, chronic kidney disease and acute kidney injury during hospital stay; indicated increased mortality post discharge. This study adds up to the accumulating data regarding prognosis and follow up of post COVID patients.

In this study, older age has been noted as an independent factor for increased mortality among COVID patients discharged from the hospital. We found that with increasing age (aOR 1.033, 95% CI 1.007 – 1.061) was associated with increased odds of mortality after being discharged from COVID-19 illness. A long-term mortality study in Estonia also noted a similar finding with increased 1 year mortality in patients older than 60 years (8). In hospital mortality of COVID-19 illness also revealed similar findings in a meta-analysis of 611,583 patients (OR 3.13, 95% CI 2.61-3.76) (9). Similar association was found in other meta-analysis and studies conducted in other countries. (10). The association between sex and mortality has not been well defined in studies in both COVID-19 hospitalized patients and discharged patients and the results have been conflicting. In the present study too, we did not find any correlation of sex with mortality.

Poor clinical status was associated with increased post COVID mortality. Poor clinical status comprised of patients having more oxygen requirement at admission and requiring higher flow rates of oxygen during hospital stay. The patients who had need of ICU admission or vasopressor need any time during the hospital stay also had increased odds of mortality (aOR 2.44, 95% CI 1.212 – 4.938). A study of 13,638 patients on follow up after COVID-19 illness, revealed increased 12-month mortality in patients having severe COVID-19 infection (HR 2.50; 95% CI 2.02, 3.09). Similar studies done in hospitalized patients with COVID-19 also reflect similar results. Lower oxygen saturation at admission had increased risk of death in hospitalized COVID-19 patients also (11). Clinical status at discharged was a significant risk factor and the patients requiring oxygen at discharge had a higher risk of post COVID mortality (aOR 2.34, 95% CI 0.971 – 5.640). Dyspnea was the only clinical symptom associated with increased risk, but it was insignificant after adjusting for other variables. A meta-analysis of 3578 patients of COVID-19 illness, presence of dyspnea was associated with 3 times increased risk of mortality (12). Similar data on post COVID mortality is however not conclusive.

Among the pre-existing comorbidities examined in the study, malignancy was found to be the independent predictor of mortality in post COVID patients. Another similar study found increased mortality in follow up patients due to malignancy, cardiovascular disease and pre-morbid respiratory disease (8). In our study we did not find a correlation of mortality with hypertension, diabetes, heart disease or chronic kidney disease. Several other studies done in hospitalized patients of COVID-19 illness have found correlations of mortality with malignancy and heart disease (13) while some have not (11).

In hospital complications can contribute to mortality in COVID-19 patients. In this study, we found AKI to be associated with increased post COVID mortality (aOR 5.60, 95% CI 2.35 – 13.37). This aspect has been analysed in studies on in hospital COVID mortality and meta analysis of 44 studies found that Acute Respiratory Distress Syndrome, Acute Kidney Injury and Acute Cardiac Injury increased the risk (14).

This is one of the first studies from India to assess the predictors of mortality among COVID-19 patients discharged from a hospital setting. It highlights the significance of follow up in COVID-19 patients. However, there are certain limitations. As is true for any retrospective study, the extraction of records lacked completeness. Drugs and their adverse effects have not been included in the study. The study participants were not divided into severity categories, however other variables like saturation at admission, length of hospital stay, need for ventilation, vasopressors or ICU admission was included.

## Conclusions

Increasing age, oxygen requirement at discharge, malignancy, ICU admission, and Acute Kidney Injury were significant predictors of mortality among COVID-19-recovered patients post discharge. The presence of these variables would warrant a close follow up for these patients in order to decrease post COVID mortality. Understanding predictors of mortality will help in ongoing care of the survivors and prioritizing patient care and preventive approaches.

### Contributors

Arjun contributed to the data collection, data analysis, and was involved in manuscript writing. BJ and PD contributed to the data collection and were involved in reviewing the draft. PKP gave the concept, interpreted analysis, critically reviewed the draft, and approved it for publication along with all authors.

### Data sharing

It will be made available to others as required upon requesting the corresponding author.

## Data Availability

All data produced in the present study are available upon reasonable request to the authors.

## Acknowledgment

None

